# SARS-CoV-2 infection induces robust, neutralizing antibody responses that are stable for at least three months

**DOI:** 10.1101/2020.07.14.20151126

**Authors:** Ania Wajnberg, Fatima Amanat, Adolfo Firpo, Deena R. Altman, Mark J. Bailey, Mayce Mansour, Meagan McMahon, Philip Meade, Damodara Rao Mendu, Kimberly Muellers, Daniel Stadlbauer, Kimberly Stone, Shirin Strohmeier, Judith Aberg, David L. Reich, Florian Krammer, Carlos Cordon-Cardo

## Abstract

SARS-CoV-2 has caused a global pandemic with millions infected and numerous fatalities. Questions regarding the robustness, functionality and longevity of the antibody response to the virus remain unanswered. Here we report that the vast majority of infected individuals with mild-to-moderate COVID-19 experience robust IgG antibody responses against the viral spike protein, based on a dataset of 19,860 individuals screened at Mount Sinai Health System in New York City. We also show that titers are stable for at least a period approximating three months, and that anti-spike binding titers significantly correlate with neutralization of authentic SARS-CoV-2. Our data suggests that more than 90% of seroconverters make detectible neutralizing antibody responses and that these titers are stable for at least the near-term future.

**One Sentence Summary:** Antibody responses induced by natural mild-to-moderate SARS-CoV-2 infection are robust, neutralizing and are stable for at least 3 months.

## Main Text

Severe acute respiratory syndrome coronavirus 2 (SARS-CoV-2) has infected millions of individuals globally and, as of July 2020, has led to the death of more than 500,000 individuals. Despite the global spread of the virus, we still lack understanding of many aspects of the humoral immune response that natural infection with SARS-CoV-2 induces (*1*). Many SARS-CoV-2 infections are mild or even asymptomatic. While the antibody responses to severe COVID-19 are relatively well characterized (*2, 3*), understanding the response in mild COVID-19 cases is of high importance, since mild and asymptomatic cases constitute the majority of infections. It will be critical to understand the robustness of the antibody response in mild cases, including its longevity and its functionality, so as to inform serosurveys, as well as to determine levels and duration of antibody titers that may be protective from reinfection.

Antibodies to SARS-CoV-2 can target many of its encoded proteins, including structural and non-structural antigens. So far, two structural proteins have been utilized as target antigens for serological assays. One of them is the abundant nucleoprotein (NP), which is found inside the virus or inside infected cells. A number of antibody-based assays targeting NP have been developed, and are being used for serological studies. However, due to the biological function of NP and the fact that it is shielded from antibodies by viral or cellular membranes, it is unlikely that NP antibodies can directly neutralize SARS-CoV-2. For SARS-CoV-1, it has been shown that vaccination with NP can induce strong antibody responses; however, they were found to be non-neutralizing (*4*). While non-neutralizing antibodies might still exert antiviral activity, for example via Fc-Fc receptor-based effector function, non-neutralizing NP antibodies have led to enhanced disease for some vaccine candidates in animal models when neutralizing antibodies were absent (*4*). The second structural protein often used as target for characterizing the immune response to SARS-CoV-2 is the spike protein. The spike is a large trimeric glycoprotein that contains the receptor binding domain (RBD), which the virus uses to dock to its cellular receptor angiotensin converting enzyme 2 (ACE2), in addition to possessing the machinery that allows fusion of viral and cellular membranes (*5, 6*). It is known from other coronaviruses as well as for SARS-CoV-2 that the spike is the main, and potentially the only target for neutralizing antibodies (*7*). Therefore, the assay used in this study to characterize the antibody response to SARS-CoV-2 is based on the trimerized, stabilized ectodomain of the spike protein (*8*). An enzyme-linked immunosorbent assay (ELISA) was initially developed in early 2020, has been extensively used in research (*9-12*), and was established in Mount Sinai’s CLIA laboratory where it received New York State Department of Health (NYSDOH) and FDA emergency use authorization (EUA) (*8, 13*). The so-called Mount Sinai ELISA antibody test has high sensitivity (92.5%) and specificity (100%) as determined with an initial validation panel of samples (S. Table 1); and a positive predictive value (PPV) of 100%, with a negative predictive value (NPV) of 99.6%.

In March 2020, Mount Sinai Health System started to screen individuals for antibodies to SARS-CoV-2 to recruit volunteers as donors for convalescent plasma therapy (*14*). Screened patients either had confirmed SARS-CoV-2 infections by PCR, or suspected disease, defined as being told by a physician that symptoms may be related to SARS-CoV-2 or exposure to someone with confirmed SARS-CoV-2 infection. The vast majority of symptomatic cases that were screened experienced mild-to-moderate disease, with less than 5% requiring emergency department evaluation or hospitalization. In addition to screening potential donors, Mount Sinai also offered the Mount Sinai ELISA antibody test to all employees within our health system on a voluntary basis. By July 2^nd^, Mount Sinai had screened 51,829 individuals using ELISAs with 19,763 individuals being positive (defined as detectible antibodies to the spike protein at a titer of 1:80 or higher) and 32,063 individuals being negative. The CLIA ELISA set up results in a discrete titer at either 1:80, 1:160, 1:320, 1:960 or ≥1:2880. We have categorized titers of 1:80 and 1:160 as low titers, 1:320 as moderate, and 1:960 and ≥1:2880 as high titers. For plasma therapy, titers of 1:320 or higher were initially deemed eligible. Of the 19,763 positive samples 505 (2.56%) had a titer of 1:80, 943 (4.77%) of 1:160, 4391 (22.23%) of 1:320, 6272 (31.75%) of 1:960 and 7641 (38.68%) of 1:2880 (**Figure 1**). Therefore, the vast majority of positive individuals have moderate to high titers of anti-spike antibodies. Of course, the argument could be made that we could be missing a number of individuals that had been infected with SARS-CoV-2 and did not produce antibodies, since many individuals included in our data set had never been tested by a nucleic acid amplification test (NAAT) for the virus. An earlier analysis performed with a smaller subset of 568 PCR-confirmed individuals using the same ELISA showed that >99% of them developed an anti-spike antibody response (*9*). In a later dataset of 2,347 patients who self-reported positive PCR, 95% of them had positive antibody titers, indicating we are not missing large numbers of patients and confirming our prior sensitivity findings. We therefore report here that the rate of individuals who do not seroconvert after SARS-CoV-2 infection is low, although such individuals may exist, and the majority of responders mount titers of 1:320 or higher.

**Figure 1.**
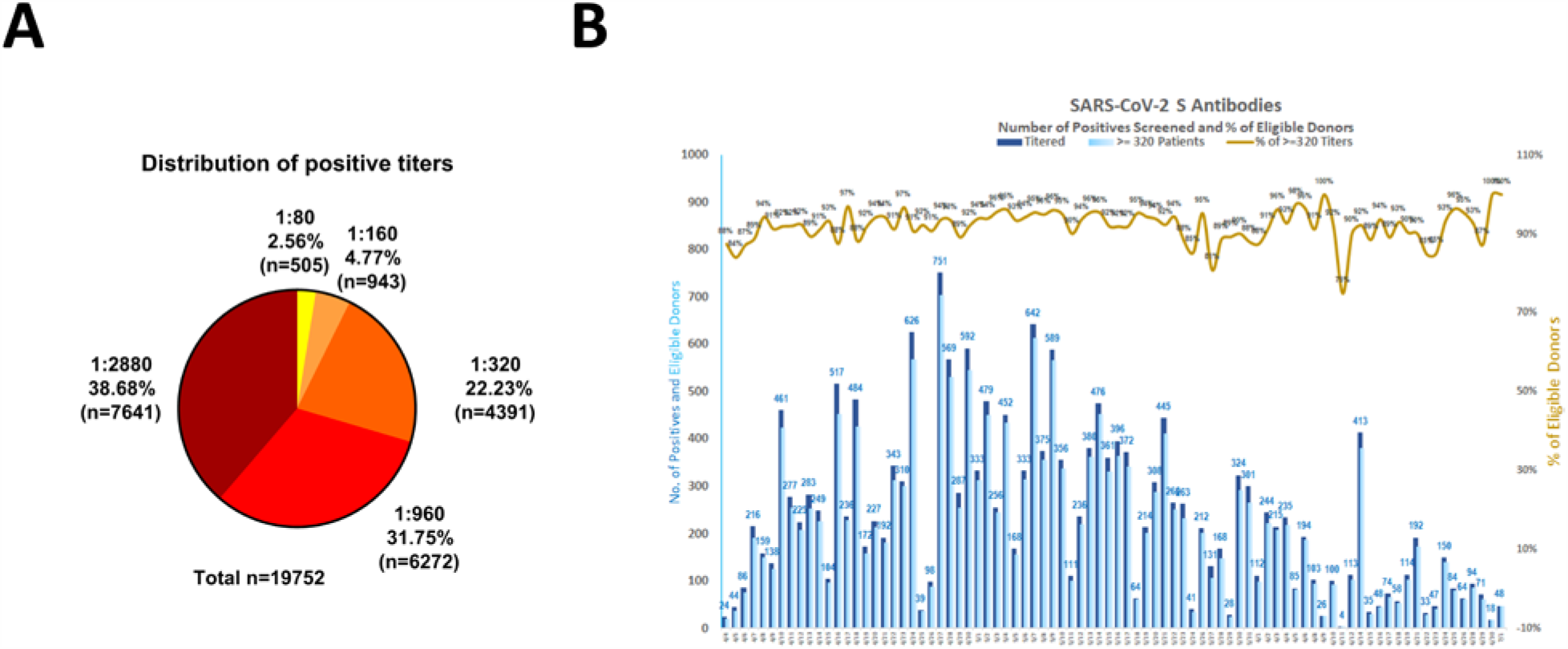
SARS-CoV-2 spike antibody titers in 19,860 individuals. **A** shows the percentage of individuals with antibody titers of 1:80 (low), 1:160 (low), 1:320 (moderate), 1:960 (high) and ≥1:2880 (high). **B** Absolute numbers and percent of individuals with titers of 1:320 over time. Over time, the screening program shifted from plasma donors (mild to moderate cases) to employee screening (with likely a higher number of asymptomatic infections) which likely caused the decrease and fluctuation in % above 1:320 at later time points.

Binding antibody titers tell us how robust the immune response to a certain virus or antigen is. However, it does not necessarily tell us anything about functionality of the antibody response, and this has been an open question related to SARS-CoV-2. Determining the neutralizing effects of SARS-CoV-2 spike antibodies is critical to understanding possible protective effects of the innate immune response. Antibodies can exert antiviral actions via many different pathways, including Fc-Fc receptor mediated effector functions and Fc-complement interactions (*15-17*). However, the antiviral activity that correlates with protection for most viruses is *in vitro* neutralization activity. In order to determine if antibodies induced against the spike protein exert neutralizing activity, we performed a well-established, quantitative microneutralization assay (*18*) based on authentic SARS-CoV-2 with 120 samples of known ELISA titers ranging from ‘negative’ to 1:2880. Neutralization titers significantly correlated (Spearman r= 0.87, p<0.0001) with spike-binding titers (**Figure 2A**). While there was some variability, sera with 1:320 ELISA titers had a geometric mean 50% inhibitory dilution (ID_50_) of approximately 1:30, for the 1:960 titer, it was approximately 1:75 and for the 1:2880 titer it was approximately 1:550. Considering any neutralizing activity above background, approximately 50% of sera in the1:80-1:160 titer range had neutralizing activity, 90% in the 1:320 range had neutralizing activity, and all sera in the 1:960-1:2880 range had neutralizing activity (**Figure 2B**). This is encouraging, and further validates our use of antibody titers to refer for plasma donation and for further investigation into protective effects at the various titer levels.

**Figure 2.**
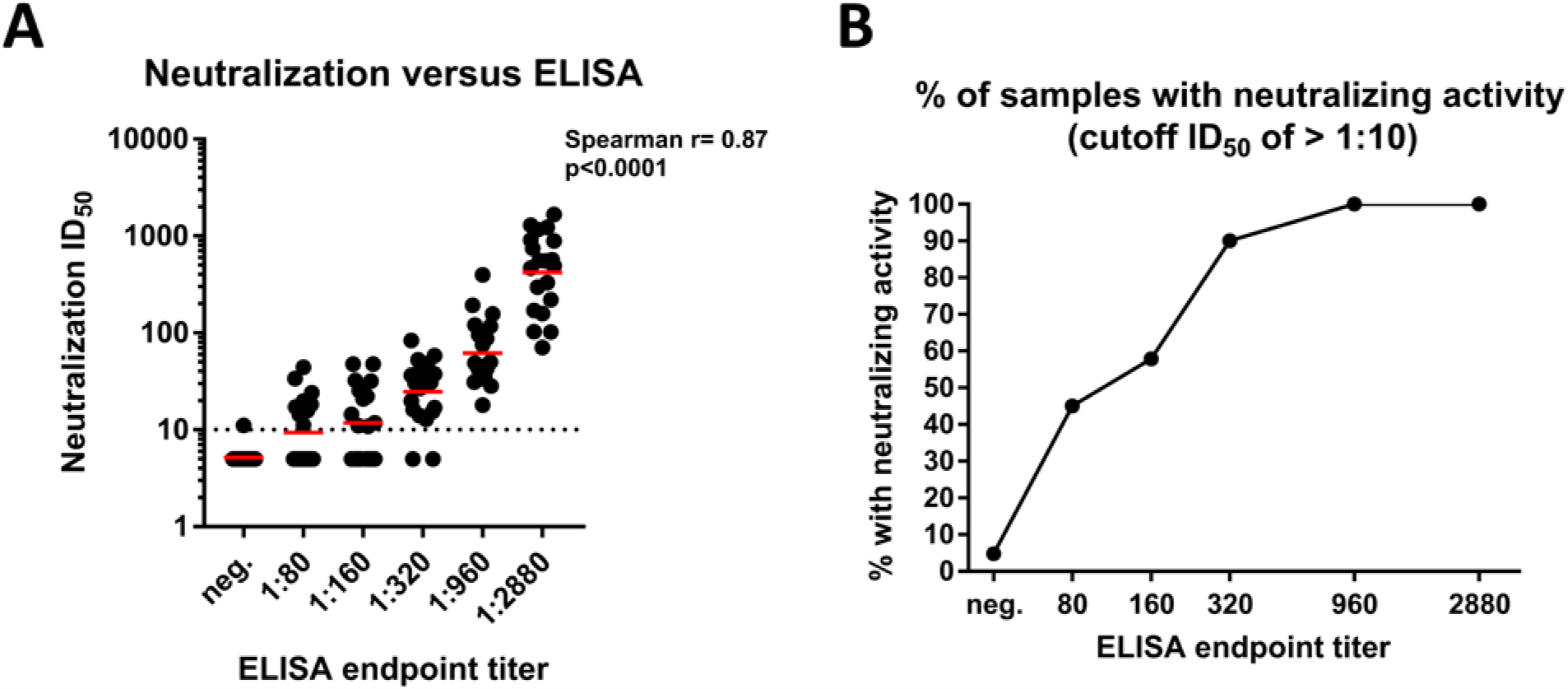
Neutralizing activity of serum samples in relation to ELISA titers. **A** shows a correlation analysis between ELISA titers on the x-axis and neutralization titers in a microneutralization assay on the y-axis. The Spearman r was determined. **B** shows the proportion of sera that exert any neutralizing activity in each of the ELISA titer categories.

Another important question is longevity of the antibody response to the spike. To assess the medium-range stability of serum antibody titers against the spike protein, we recalled 121 plasma donors at a variety of titer levels that had initially been screened at approximately day 30 post symptom onset. The mean interval between the initial titer measurement and the second was 52 days (range 33-67 days), setting the second time point at a mean of 82 days post symptom onset (range 52-104 days). When we compared overall titers we saw a slight drop from a geometric mean titer (GMT) of 670 to a GMT of 642 (**Figure 3A**). In the higher titer range of 1:2880 and 1:960, we also observed a slight drop in titer between the two time points (**Figure 3B and C**). Surprisingly, but in agreement with earlier observations from our group that seroconversion in mild COVID-19 cases might take longer time to mount (*9*), we saw an increase in individuals who had an initial titer of 1:320, 1:160 or 1:80 (**Figure 3D-F**). The exception was one individual that dropped from a 1:80 titer to being negative. The initial serum antibody titer was likely produced by plasmablasts, and plasmablast-produced antibody peaks 2-3 weeks post symptom onset. Given an IgG half-life of approximately 21 days, the sustained or increasing antibody titers observed are likely produced by long-lived plasma cells in the bone marrow. Of note, our observations are in contrast to a recent report by Long *et al*. that found waning titers 8 weeks post virus infection as compared to acute responses (*19*). Especially in asymptomatic cases, antibody responses disappeared after 8 weeks in 40% of individuals in the Long *et al*. study. However, the antibodies measured in their paper were targeting the NP plus a single linear spike epitope. Much more in agreement with our data, the same paper also reports relatively stable (slightly declining) neutralizing antibody titers. The stability of the antibody response over time might therefore also depend on the target antigen. The titers we measure here do correlate with neutralization as discussed above.

**Figure 3.**
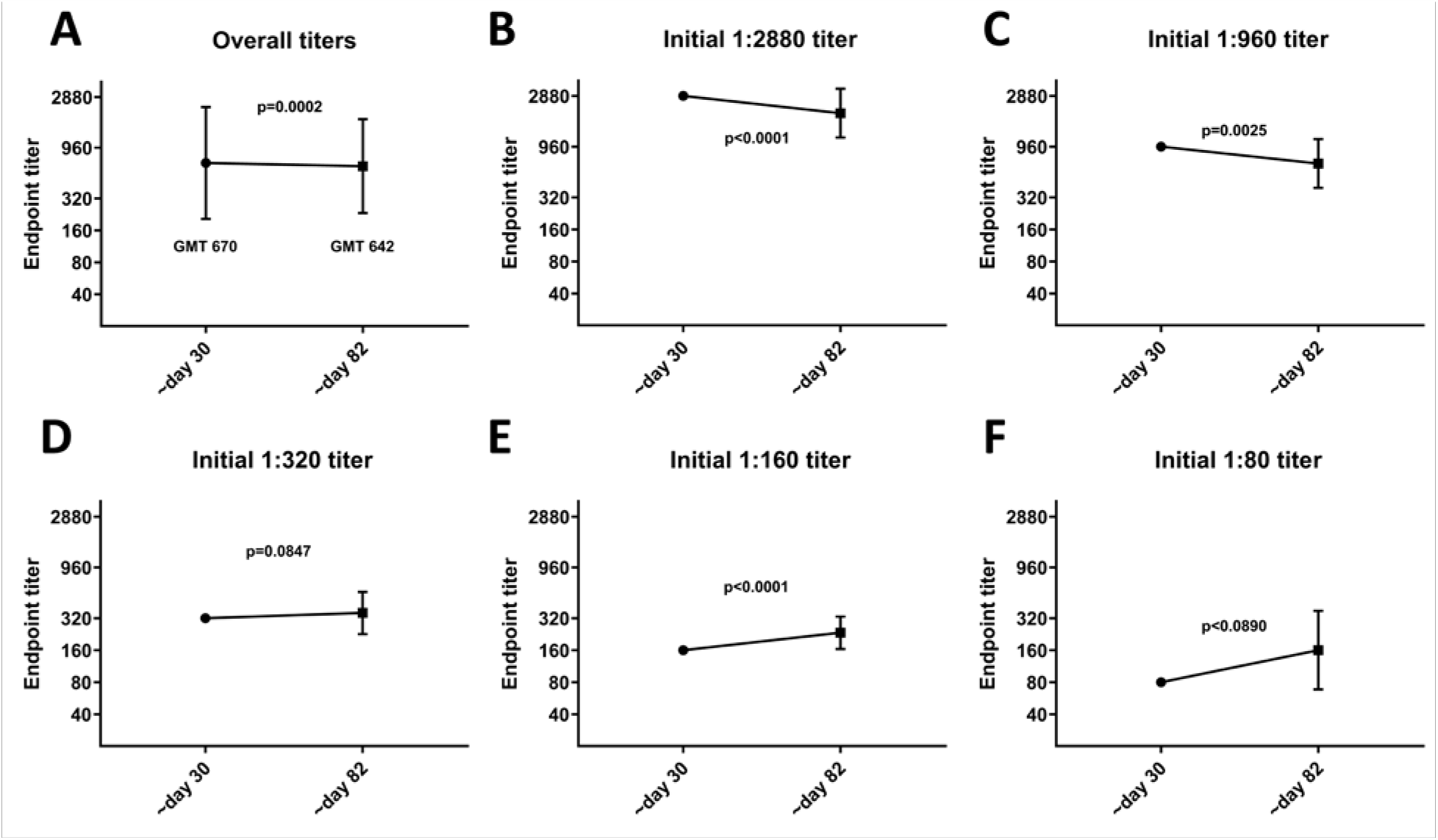
Antibody titer stability over time. **A** shows titers of 121 volunteers who were initially bled approximately 30 days post COVID-19 symptom onset and were then recalled and bled again approximately 82 days post symptom onset. **B, C, D, E and F** shows the same data but stratified by initial/day 30 titer. Titers are graphed as geometric mean titers (GMT).

For many different viral infections correlates of protection have been established. These correlates are usually based on a specific level of antibody acquired through vaccination or natural infection that significantly reduces the risk of (re-)infection. Examples are the hemagglutination inhibition titer for influenza virus, where a 1:40 titer reduces the risk of getting infected by 50% (*20*). Similar titers have been established for measles virus (an ID_50_ titer of 1:120), hepatitis A virus, hepatitis B virus and many others (*21*). These titers have facilitated vaccine development significantly. For some viruses/vaccines, the kinetics of the antibody response is also known, allowing for an accurate prediction of how long protection will last (*22*).

It is still not clear if infection with SARS-CoV-2 in humans protects from re-infection and for how long. We know from work with common human coronaviruses that neutralizing antibodies are induced and these antibodies can last for years and provide protection from re-infection or attenuate disease, even if individuals get re-infected (*23*). Furthermore, we now know from non-human primate models that infection with SARS-CoV-2 does protect from re-infection for at least some time (*24, 25*). This protection is, in some cases, more pronounced in the lower respiratory tract than in the upper respiratory tract (*25*). We also know that transferring serum, for example from infected hamsters into naïve hamsters, reduces virus replication significantly when the naïve hamsters are challenged (*26*). Similarly, non-human primates can be protected by prophylactic treatment with neutralizing monoclonal antibodies (*27*). Finally, vaccine induced neutralizing antibody titers have been established as correlate of protection in non-human primates (*28*). Of note, these titers were relatively low and in the lower range of the titers observed here. Our data reveal that individuals who have recovered from mild COVID-19 experience robust antibody responses. Antibody binding titers to the spike protein correlate significantly with neutralization with authentic SARS-CoV-2 virus, and the vast majority of individuals with antibody titers of 1:320 or higher show neutralizing activity in their serum. Consistent with data for human coronaviruses, SARS-CoV-1 and Middle Eastern respiratory syndrome-CoV (*23*), we also find stable antibody titers over a period of at least 3 months, and we plan to follow this cohort over longer intervals of time. While this cannot provide conclusive evidence that these antibody responses protect from re-infection, we believe it is very likely that they will decrease the odds ratio of getting re-infected, and may attenuate disease in the case of breakthrough infection. We believe it is imperative to swiftly perform studies to investigate and establish a correlate of protection from infection with SARS-CoV-2. A correlate of protection, combined with a better understanding of antibody kinetics to the spike protein, would inform policy regarding the COVID-19 pandemic and would be of benefit to vaccine development.

## Data Availability

Raw data is available from the corresponding authors upon reasonable request.

## Acknowledgements

We are grateful for the continuous expert guidance provided by the ISMMS Program for the Protection of Human Subjects (PPHS). We also thank Dr. Randy A. Albrecht for oversight of the conventional BSL3 biocontainment facility, the medical students involved in the plasma convalescence program. Furthermore, we would like to express our gratitude to Dr. Erik Lium and team at Mount Sinai Innovation Partners for continuous support and to Vanesa Sarić and her team at Mount Sinai’s Development Office for fundraising and for taking many little things off our shoulders during this difficult time. Finally, we would like to thank Dr. Dennis Charney and the Dean’s Office for strong institutional support of our work. This work was partially supported by the NIAID Centers of Excellence for Influenza Research and Surveillance (CEIRS) contract HHSN272201400008C (FK), Collaborative Influenza Vaccine Innovation Centers (CIVIC) contract 75N93019C00051 (FK), and the generous support of the JPB foundation, the Open Philanthropy Project (#2020-215611) and other philanthropic donations.

## Conflict of interest statement

Mount Sinai has licensed serological assays to commercial entities and has filed for patent protection for serological assays.

**Supplemental Table 1:**
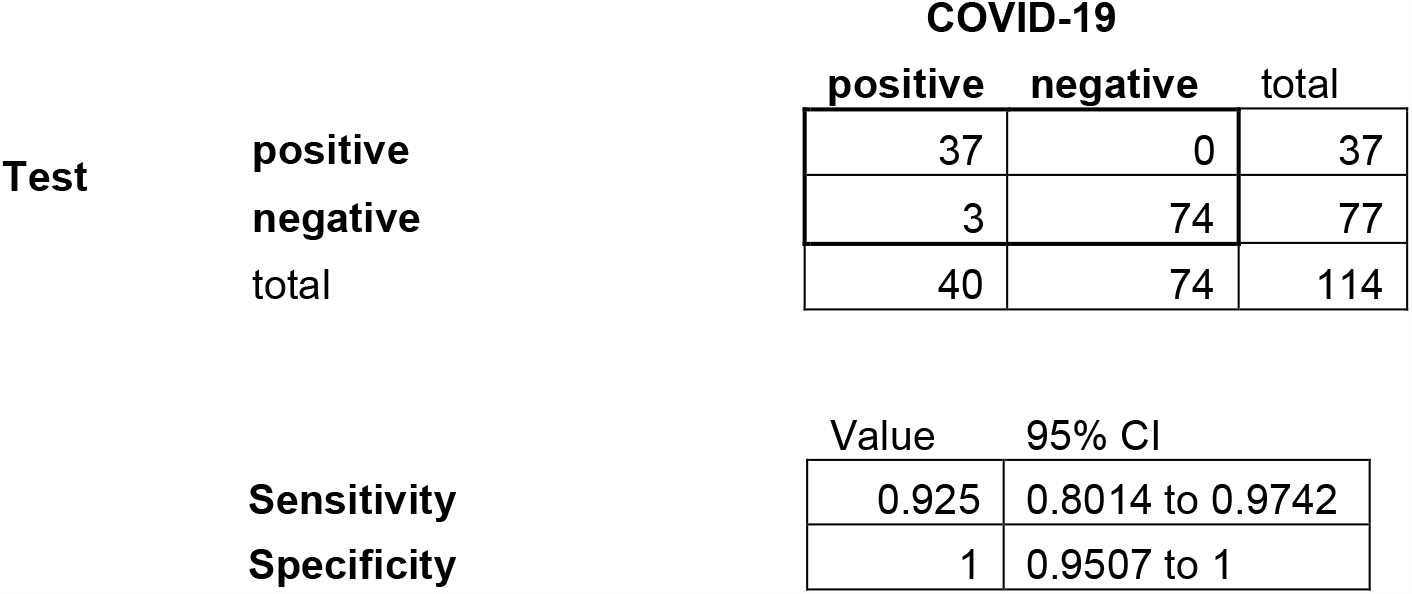
**Contingency table to calculate sensitivity and specificity of the Mount Sinai ELISA SARS-CoV-2 antibody test**

| Test |  | COVID-19 |  |  |
| --- | --- | --- | --- | --- |
|  |  | positive | negative | total |
|  | positive | 37 | 0 | 37 |
|  | negative | 3 | 74 | 77 |
|  | total | 40 | 74 | 114 |

|  | Value | 95% CI |
| --- | --- | --- |
| Sensitivity | 0.925 | 0.8014 to 0.9742 |
| Specificity | 1 | 0.9507 to 1 |

## Materials and Methods

### Study participants, human samples and study design

Starting in late March, 2020, we conducted an outreach program in the New York City area, including parts of New York State, Connecticut, and New Jersey, to identify people recovered from SARS-CoV-2. Participants were tested for SARS-CoV-2 antibodies and, if positive, obtained a titer level (1:80, 160, 320, 960 or ≥2880) using the Mount Sinai ELISA described here. We recruited participants via REDCap^®^ (Vanderbilt University, Tennessee) online survey response which was advertised on our hospital website, and subsequently shared by multiple news organizations and public officials in New York. REDCap respondents’ were deemed eligible if they had previously tested positive for SARS-CoV-2 via nasopharyngeal PCR (cobas^®^ SARS-CoV-2, Roche Diagnostics, Indiana). Due to the lack of PCR testing in the New York area prior to mid-March, 2020, we also included people for antibody testing and screening if they were symptomatic with suspected SARS-CoV-2 symptoms after February 1, 2020, if they had a high risk exposure to someone with a positive SARS-CoV-2 PCR test, or were healthcare workers. Additionally, only participants who were asymptomatic at time of survey were eligible to come in for antibody testing. Respondents self-reported date of symptom onset, date of positive SARS-CoV-2 test (if applicable), and last date of symptoms.

During the first week of serum antibody testing, participants were brought in ten or more days after they had a confirmed/suspected diagnosis and had been asymptomatic for at least three days. In week 2, as we identified more potential donors and learned more about our antibody assay, we extended our timeline to 14 days after symptoms onset, with at least three days asymptomatic. We saw quickly that this was a little early for serum conversion, and by week 3, we included participants 21 days or more after symptom onset, who had been completely asymptomatic for at least 14 days. We referred donors for plasma donation to the New York Blood Center if their antibodies were ≥1:320. All interested participants with antibody titers >1:320 and negative SARS-CoV-2 PCR swabs were screened by the New York Blood Center using standard criteria for plasma donation and included as donors in our convalescent plasma study if eligible as per CFR Title 21. Studies were reviewed and approved by our institutional review board.

It is important to note that as we identified greater numbers of potential donors and built capacity for testing in our clinical lab, we offered testing to wider groups including family and friends of indivduals with positive PCR or antibodies, essential workers, patients in our health system, members of our community, and finally to all employees of the Mount Sinai Health System. Testing for employees was voluntary, and included both front line health care workers and administrative support staff, faculty and students, regardless of their clinical assignments during the COVID-19 pandemic. This broadening likely explains why our antibody testing yielded lower positive rates over time, not because of characteristics of the test or of the population curve.

### Enzyme-linked immunosorbent assay (ELISA)

The Mount Sinai Hospital (MSH) enzyme linked immunosorbent assay (ELISA) is an orthogonal immune assay specific for IgG anti SARS-CoV-2 spike protein in serum or plasma and measures the relative concentration of IgG as the highest dilution of serum giving a positive signal (OD_490_ ≥ 0.15) after exactly 5 minutes of color development. The result is reported as reciprocal value of the sample’s highest dilution producing a signal. The assay received FDA authorization for clinical use on 4/15/202 validating the safe application on the original research assay developed and described by the Krammer laboratory team in the CLIA environment. The MSH-ELISA Anti IgG COVID-19 assay was also independently authorized as a laboratory developed test (LDT) for clinical application by the NYSDOH at the Mount Sinai Laboratory (MSL), Center for Clinical Laboratories, a division of the Department of Pathology, Molecular, and Cell-Based Medicine, New York, NY (CLIA# 33D1051889) from individuals suspected of previous COVID-19 infection by their healthcare provider, for the assessment of seroconversion from an antibody negative status to an antibody positive status in acutely infected patients, and for identification of individuals with SARS-Cov-2 IgG antibodies titers of up to 1:2880. A detailed description of the assay can be found here (*8, 13*).

### Neutralization assay

Human samples were heat-inactivated at 56°C for an hour prior to use. Vero.E6 cells (ATCC #CRL-1586) were seeded at a density of 20,000 cells per well in a 96-well cell culture plate (Corning, cat. no. 3595) one day before the assay was performed. Cells were maintained in culture in complete Dulbecco’s Modified Eagle’s Medium and the media used for the neutralization assay was 1X Minimal Essential Medium supplemented with 2% fetal bovine serum (FBS; Corning). The details of the protocol have been described in detail earlier (*18*).

Starting with 1:10, serial dilutions of each sample was prepared in a 96-well plate in duplicates. Six wells in each plate were used as “no virus” controls and 6 wells were used as “virus only” controls. Next, 80uls of each respective dilution was mixed with 600TCID_50_ of SARS-CoV-2 isolate USA-WA1/2020, (BEI Resources NR-52281) in 80 uls. The virus-serum mixture was incubated for an hour. Next, media from cells was removed and 120 uls of virus-serum mixture was added onto the cells. The cells were incubated for an hour at 37°C. After an hour, the virus-serum mixture was removed and 100 uls of 1X MEM and 100 uls of each respective serum dilution was added to the cells. The cells were kept in the 37°C incubator for 2 days. After 48 hours, the media from the cells was removed and 150 uls of 10% formaldehyde (Polysciences) was added to the cells to inactivate the virus for 24 hours. The next day, cells were permeabilized and stained using an anti-NP antibody. Percent inhibition of virus was calculated for each dilution. This protocol has been earlier published in much greater detail (*18*).

### Statistical analysis

Correlation analysis was performed using Spearman’s rank test. A paired t-test was used to compare longitudinal titers. A p>0.05 was considered significant. Analysis was performed in GraphPad Prism.

## References and Notes

1. F. Krammer, V. Simon, Serology assays to manage COVID-19. Science 368, 1060–1061 (2020).

2. L. Liu et al., High neutralizing antibody titer in intensive care unit patients with COVID-19. Emerg Microbes Infect, 1–30 (2020).

3. Q. X. Long et al., Antibody responses to SARS-CoV-2 in patients with COVID-19. Nat Med 26, 845–848 (2020).

4. D. Deming et al., Vaccine efficacy in senescent mice challenged with recombinant SARS-CoV bearing epidemic and zoonotic spike variants. PLoS Med 3, e525 (2006).

5. D. Wrapp et al., Cryo-EM structure of the 2019-nCoV spike in the prefusion conformation. Science, (2020).

6. M. Letko, A. Marzi, V. Munster, Functional assessment of cell entry and receptor usage for SARS-CoV-2 and other lineage B betacoronaviruses. Nat Microbiol 5, 562–569 (2020).

7. F. Amanat, F. Krammer, SARS-CoV-2 Vaccines: Status Report. Immunity 52, 583–589 (2020).

8. F. Amanat et al., A serological assay to detect SARS-CoV-2 seroconversion in humans. Nat Med, (2020).

9. A. Wajnberg et al., Humoral immune response and prolonged PCR positivity in a cohort of 1343 SARS-CoV 2 patients in the New York City region. medRxiv, 2020.2004.2030.20085613 (2020).

10. A. S. Dingens et al., Seroprevalence of SARS-CoV-2 among children visiting a hospital during the initial Seattle outbreak. medRxiv, (2020).

11. D. S. Hains et al., Asymptomatic Seroconversion of Immunoglobulins to SARS-CoV-2 in a Pediatric Dialysis Unit. JAMA, (2020).

12. D. Stadlbauer et al., Seroconversion of a city: Longitudinal monitoring of SARS-CoV-2 seroprevalence in New York City. medRxiv, 2020.2006.2028.20142190 (2020).

13. D. Stadlbauer et al., SARS-CoV-2 Seroconversion in Humans: A Detailed Protocol for a Serological Assay, Antigen Production, and Test Setup. Curr Protoc Microbiol 57, e100 (2020).

14. S. T. H. Liu et al., Convalescent plasma treatment of severe COVID-19: A matched control study. medRxiv, 2020.2005.2020.20102236 (2020).

15. N. K. Thulin, T. T. Wang, The Role of Fc Gamma Receptors in Broad Protection against Influenza Viruses. Vaccines (Basel) 6, (2018).

16. B. M. Gunn et al., A Role for Fc Function in Therapeutic Monoclonal Antibody-Mediated Protection against Ebola Virus. Cell Host Microbe 24, 221-233.e225 (2018).

17. E. O. Saphire, S. L. Schendel, B. M. Gunn, J. C. Milligan, G. Alter, Antibody-mediated protection against Ebola virus. Nat Immunol 19, 1169–1178 (2018).

18. F. Amanat et al., An In Vitro Microneutralization Assay for SARS-CoV-2 Serology and Drug Screening. Curr Protoc Microbiol 58, e108 (2020).

19. Q. X. Long et al., Clinical and immunological assessment of asymptomatic SARS-CoV-2 infections. Nat Med, (2020).

20. F. Krammer, J. P. Weir, O. Engelhardt, J. M. Katz, R. J. Cox, Meeting report and review: Immunological assays and correlates of protection for next-generation influenza vaccines. Influenza Other Respir Viruses, (2019).

21. S. A. Plotkin, Correlates of protection induced by vaccination. Clin Vaccine Immunol 17, 1055–1065 (2010).

22. K. Van Herck, P. Van Damme, Inactivated hepatitis A vaccine-induced antibodies: follow-up and estimates of long-term persistence. J Med Virol 63, 1–7 (2001).

23. A. T. Huang et al., A systematic review of antibody mediated immunity to coronaviruses: antibody kinetics, correlates of protection, and association of antibody responses with severity of disease. medRxiv, 2020.2004.2014.20065771 (2020).

24. W. Deng et al., Primary exposure to SARS-CoV-2 protects against reinfection in rhesus macaques. Science, (2020).

25. A. Chandrashekar et al., SARS-CoV-2 infection protects against rechallenge in rhesus macaques. Science, (2020).

26. M. Imai et al., Syrian hamsters as a small animal model for SARS-CoV-2 infection and countermeasure development. Proc Natl Acad Sci U S A, (2020).

27. R. Shi et al., A human neutralizing antibody targets the receptor binding site of SARS-CoV-2. Nature, (2020).

28. J. Yu et al., DNA vaccine protection against SARS-CoV-2 in rhesus macaques. Science, (2020).

